# Converting Microwave Ovens into Plasma-Generating Decontamination Units for N-95 Respirators

**DOI:** 10.1101/2020.07.28.20163915

**Authors:** David N. Ruzic, Chamteut Oh, Joseph V. Puthussery, Vishal Verma, Thanh H. Nguyen

## Abstract

We show how a common microwave oven, a coat-hanger and a coffee cup can be used to decontaminate N-95 respirators in 30 seconds. Tulane virus in artificial saliva was reduced by >3 log and *Geobacillus stearothermophilus* spores were reduced by >6 log. Respirators maintained filtration and fitting after 10 cycles.

## Text

The ongoing pandemic of SARS-CoV-2 (i.e. COVID-19) has severely stressed the worldwide healthcare system and has created unprecedented shortages of personal protective equipment (PPE) including N-95 filtering respirators (N-95s). Plasma sterilization is a known technique and works on viruses [1]. A factor of 30 reduction of human norovirus was obtained after a 5-minute treatment from a cold atmospheric-pressure air plasma by a FlatPlasTer 2.0™ device. The inactivation of the virus particles’ functions occur through synergistic effects of the cold plasma-initiated air chemistry [1] and UV light. [2]

Commercial plasma sterilization devices, are not widely available and are costly. Few if any hospitals, nursing homes, or medical facilities have such a device. However, everyone has a microwave oven – often on every floor. Microwave ovens have been used to make plasmas. There are numerous how-to videos using grapes on the internet [3] and scholarly articles explaining how they work [4]. Our lab specializes in using microwave sources to make atmospheric-pressure plasmas too for a variety of uses [5]. The objective of this study was to demonstrate that an atmospheric-pressure plasma generated by the microwave oven can decontaminate the respirator. We proved that 30 s of the plasma treatment can decontaminate the respirator (i.e., >3-log_10_ virucidal and >6-log_10_ sporicidal efficacy) without compromising the respirator integrity (i.e., filtration efficiency and fit testing). See Table 1.

**Table 1.** Summary of the microwave oven experiments with different experiment conditions.

| Conditions | Spore test | Virus $\log_{10}$ reduction | | Filtration efficiency (%) | Fit score |
| --- | --- | --- | --- | --- | --- |
|  |  | Inside respirator | Outside respirator |  |  |
| Control | NA<sup>a</sup> | 0 | 0 | 99.4 | NA |
| Microwave (60 s) | Fail | $0.29 \pm 0.12^b$ | $-0.08 \pm 0.17^b$ | 99.6 | NA |
| Microwave with H<sub>2</sub>O<sub>2</sub> and saline (60 s) | Fail | $0.9^c$ | $4.08^{c,e}$ | 99.5 | NA |
| Plasma (30 s) | Pass | $4.22 \pm 0.40^{d,e}$ | $4.22 \pm 0.79^{d,e}$ | 99.5 | $115 \pm 23$ |

a) Not applicable.
b) Microwave alone was applied to 5 mm diameter coupons of respirator (N=3).
c) Microwave with H<sub>2</sub>O<sub>2</sub> and saline experiment were done with the 5 mm diameter coupons of respirator (N=1).
d) Plasma treatment was performed with the whole N95 respirator with virus inoculation on different locations of the respirator (N=5).
e) The actual inactivation efficacy could be higher than the presented value because of the detection limit. The detection limit of inactivation efficacy was determined by the ratio of initial viral infectivity to the viral infectivity after treatment. Initial virus concentrations were little different and thus the detection limit was also different.

The new idea described in this work is to create a plasma by making an antenna that has a length equal to some multiples of the wavelength of the microwaves (12.2 cm) in a microwave oven bent into a split ring with a 2mm gap. An intense electric field is generated in the gap and can ignite a plasma if a conducting liquid, such as saline solution, is added to the gap. The conducting fluid electrically shorts the gap causing a large current to flow and heats the fluid quickly. The fluid is vaporized in a few seconds and a spark is generated in the gap. These first electrons from the sparks allow a plasma to be generated out of the air and other gasses present. This plasma is sustained as long as the microwave power is present. Hydrogen peroxide (30%) is also added to the gap with the saline. This plasma produces hydrogen peroxide vapor, OH radicals, UV light, O radicals, positive and negative ions, ozone, NO compounds, and electrons. All of these species are biologically active and react away any mucus/saliva barriers and then destroy the cell walls of the biological contaminant.

The ability to turn a microwave oven into a decontamination device with common everyday materials such as a coat-hanger, coffee cup and some fluids means instant world-wide accessibility. Hospitals, nursing homes, meat-packing plants, etc., could practice this technique with materials at hand and perform decontaminations in as little as 30 seconds.

The procedure to do this process is as follows: 1) Obtain a piece of heavy gauge wire which is stripped of insulation, such as a metal coat hanger. Cut it to 24.4 cm. Bend it into a circle leaving a gap of around 2mm. (Through experimentation the length must be within 5% ie. 24 to 25 cm and the gap must be between 1 and 2.5 mm.) 2) Disable the rotation in the microwave oven. This can be simply done by turning the turn-table dish upside down. 3) Place the antenna with the gap down into a shallow glass or ceramic dish. An inverted coffee cup works well. Both ends of the antenna forming the gap should be touching the ceramic/glass surface. The antenna may need to be tilted up and supported. See Figure 1. 4) Add saline solution (around 1 ml) and 30% hydrogen peroxide (at least 1ml) in the antenna gap. 5) Turn the microwave on its highest power (normal operation) for 1 minute maximum. At first the liquid will start boiling. In about 15 seconds a popping sound and bright sparks will emanate from the gap. Soon, a continuous plasma will form, and a loud buzzing sound will be heard. The continuous plasma should run for at least 30 seconds. Do not leave the mask in for more than 1 minute. If the mask is left in too long (about 2 minutes) the mask material around the metal nose piece may start to melt.

**Figure 1.**
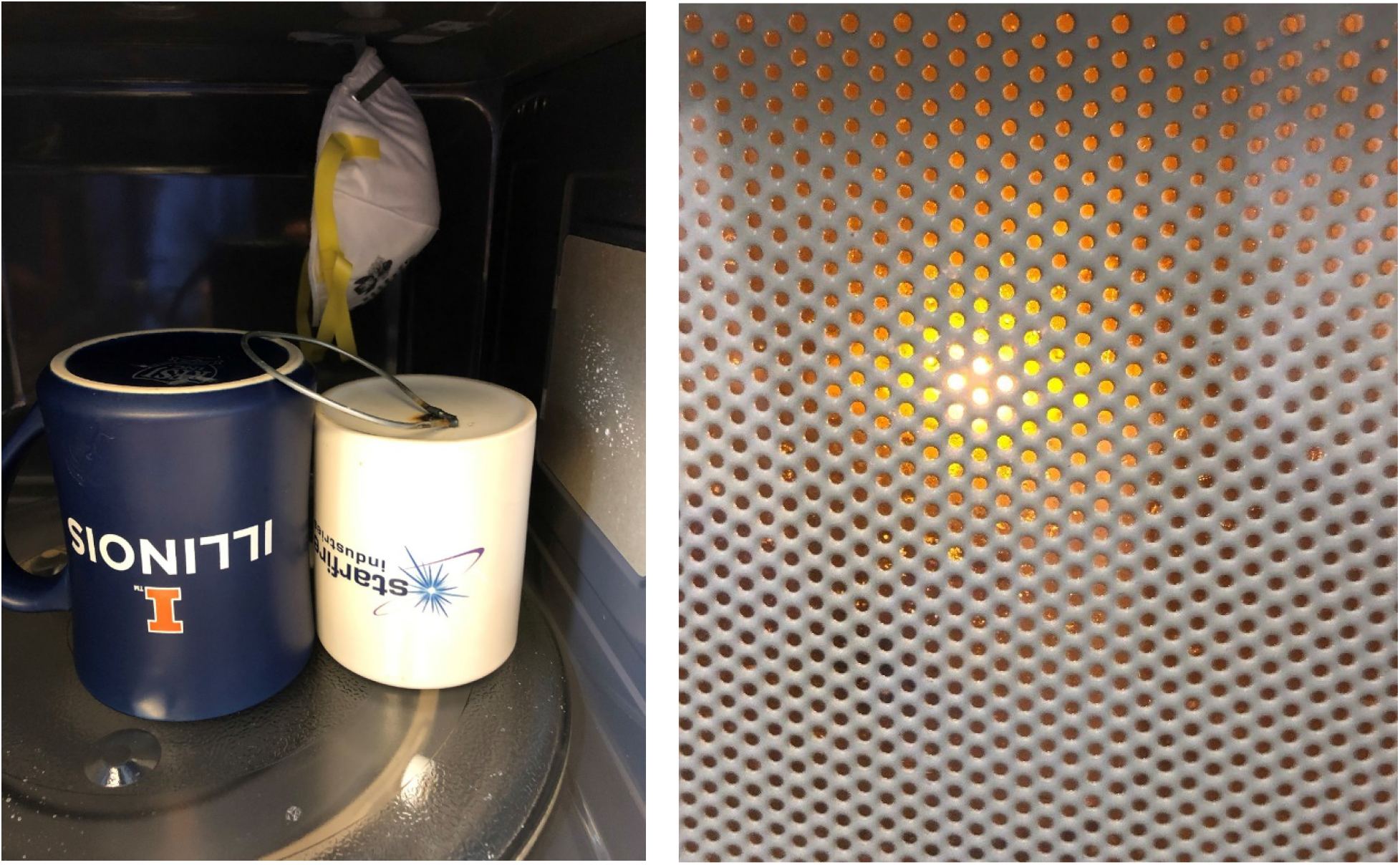
Left: antenna on inverted coffee cup in microwave oven with N-95 respirator suspended behind it. Right: a picture of the intense plasma created at the antenna gap on the cup surface of the white cup.

A few notes of caution. First, if possible one should dispose of contaminated masks and use new ones to assure optimal safety. If this procedure is used to decontaminate N-95 respirators, it is important that an actual plasma is made. It is quite obvious when the system goes into the continuous mode: A bright light is emitted and a loud buzzing sound is made by the plasma. The antenna gets very hot – so hot that the ends glow. It should not be touched until it cools down. If plastic or other flammable material touches the red-hot glowing antenna, it may catch on fire. This is likely why microwave manufacturers do not recommend that people put metal into microwave ovens! As shown by our results however, the plasma species generated in this manner are capable of decontamination in 30 seconds and anyone with a microwave oven and a few simple household items can create a N-95 respirator decontamination unit for emergency use.

## Data Availability

All data is in laboratory notebooks, and stored in computer files, of the authors

## Acknowledgements

Funding for this work was provided by the JUMP-ARCHES program of OSF Healthcare in conjunction with the University of Illinois.

## Author Bio

David Ruzic is the Able Bliss Professor in Nuclear, Plasma and Radiological Engineering and holds a zero-time appointment in the Carle-Illinois College of Medicine. He creates and studies plasma sources for use in microelectronic processing and fusion energy.

